# Offspring cardiometabolic outcomes and postnatal growth trajectories after exposure to maternal SARS-CoV-2 infection

**DOI:** 10.1101/2023.08.16.23294170

**Authors:** Lydia L. Shook, Victor M. Castro, Emily M. Herzberg, Lindsay T. Fourman, Anjali J. Kaimal, Roy H. Perlis, Andrea G. Edlow

## Abstract

**Context:** Prior birth cohorts have suggested an association between maternal infection in pregnancy and offspring risk for childhood obesity. Whether maternal SARS-CoV-2 infection is similarly associated with increased cardiometabolic risk for offspring is not known.

**Objective:** To determine whether in utero exposure to SARS-CoV-2 is associated with increased risk for cardiometabolic diagnoses by 18 months after birth, compared with unexposed offspring born during the COVID-19 pandemic.

**Design:** This retrospective cohort study included the live offspring of all individuals who delivered during the COVID-19 pandemic (April 1, 2020 - December 31, 2021) at 8 hospitals within 2 health systems in Massachusetts.

**Exposure:** SARS-CoV-2 positivity on polymerase chain reaction (PCR) test during pregnancy.

**Main Outcome Measures:** Electronic health record documentation of *International Statistical Classification of Diseases and Related Health Problems, Tenth Revision* diagnostic codes corresponding to cardiometabolic disorders. Offspring weight-for-age, length-for-age, and body mass index (BMI)-for-age z-scores at birth, 6 months, 12 months, and 18 months of age.

**Results:** The full study cohort includes 29,510 live born offspring (1,599 exposed and 27,911 unexposed offspring). 6.7% of exposed and 4.4% of unexposed offspring had received a cardiometabolic diagnosis by 18 months of age (crude OR 1.47 [95% CI: 1.10-1.94], p=0.007; adjusted OR 1.37 [1.01-1.83]; p=0.04). These diagnoses were preceded by significantly greater mean BMI-for-age z-scores in exposed versus unexposed offspring at 6 months (mean z-score difference 0.19, 95% CI: 0.10, 0.29, p<0.001), and a greater proportion of offspring at risk of, or meeting criteria for, overweight/obesity (16.5% vs. 12.2%, p=0.006).

**Conclusions:** Exposure to maternal SARS-CoV-2 infection was associated with an increased risk of receiving a cardiometabolic diagnosis by 18 months and greater BMI-for-age at 6 months.

## Introduction

Prenatal exposures can have a lasting impact on the health of the developing fetus. Psychosocial stress, smoking, alcohol consumption, obesity, and poor maternal nutrition during pregnancy have all been associated with offspring risk for obesity and cardiometabolic disease later in life (1–5). The direct impact of maternal infection and immune activation on offspring cardiometabolic outcomes has received relatively less attention. An early cross-sectional cohort study of adult male Danish military conscripts first pointed to the association between severe maternal infection and offspring obesity in adulthood, reporting that men were 34% more likely to have obesity if their mothers were hospitalized with infection during pregnancy (6). Several subsequent birth cohort studies identified modest associations between acute and chronic maternal infections and offspring obesity (7–10). Studies using animal models of infection during pregnancy have been useful in further identifying mechanisms of cardiometabolic programming (11) that could explain these effects. These investigations suggest that, as has been repeatedly shown in the fetal brain, maternal-placental immune activation itself may be an important final common pathway in the programming of cardiometabolic organs and tissues in the developing fetus (12).

SARS-CoV-2 infection during pregnancy can generate significant maternal immune responses that may have implications for the long-term health of offspring (13). As of the end of July 25, 2022, more than 225,000 cases of SARS-CoV-2 during pregnancy had been reported to the Centers for Disease Control and Prevention (CDC) (14), a figure which has likely risen substantially in the year since with the widespread availability of rapid home antigen testing and high transmissibility of the Omicron variant. Given the significant number of impacted pregnancies, even relatively small increases in chronic disease risk could be impactful from a public health perspective.

Our group has previously reported an association between in utero exposure to SARS-CoV-2 and risk of neurodevelopmental diagnoses in children at one year of age (15) and a sex bias favoring diagnoses in male children in a larger cohort extended to 18 months of follow-up (16). More recently, in a small prospectively enrolled cohort of 276 infants born during the SARS-CoV-2 pandemic identified from a COVID-19 biobank, we found evidence of accelerated postnatal weight gain in the first 12 months of life in infants whose mothers had SARS-CoV-2 during pregnancy (17), a growth pattern that may be associated with childhood obesity and cardiometabolic risk (18–20). Here, we assembled a large electronic health record (EHR)-based pregnancy cohort with linked offspring follow-up data, to evaluate the association between maternal SARS-CoV-2 infection during pregnancy and subsequent risk of a cardiometabolic diagnosis in offspring through the first 18 months of life. We next assessed differences in growth trajectories in SARS-CoV-2 exposed and unexposed offspring in this large cohort, as defined by weight, length, and BMI z-scores from birth to 18 months of age.

## Materials and Methods

### Study design and data set generation

We queried the EHR of 8 hospitals and all affiliated outpatient networks within the Mass General Brigham (MGB) system sharing a common electronic data warehouse and governance. Data were extracted from 2 academic medical centers [Massachusetts General Hospital, Brigham and Women’s Hospital] and 6 community hospitals [Newton-Wellesley Hospital, Salem Hospital, Martha’s Vineyard Hospital, Nantucket Cottage Hospital, Cooley Dickinson Hospital, and Wentworth Douglass Hospital]. All live births occurring between April 1, 2020 and December 31, 2021 at these hospitals were identified and comprised the study cohort. We linked offspring to the birthing person based upon medical record number, date and time of birth, and offspring sex. This approach to cohort definition has been successfully employed and validated by our group in prior work (15,16,21).

For birthing persons, we queried *International Statistical Classification of Diseases and Related Health Problems, Tenth Revision* (ICD-10) billing codes, problem lists, medications, and laboratory studies occurring from date of estimated last menstrual period up to the discharge date of the delivery admission. Sociodemographic factors were extracted from the EHR including maternal age, self-reported race and ethnicity based on US census categories, insurance type (public or private payer), and delivery hospital type (academic medical center or community hospital). Offspring sex, preterm birth, pre-existing or gestational diabetes (defined by ICD-10 code bundle CCS 186, “Diabetes or abnormal glucose tolerance complicating pregnancy; childbirth; or the puerperium”), maternal pre-pregnancy body mass index (BMI), and date of SARS-CoV-2 positive PCR test were also extracted. Maternal pre-pregnancy BMI was defined as closest BMI reported in the medical record relative to last menstrual period (LMP), from 6 months prior to LMP up to 2 weeks after LMP. If no pre-pregnancy value was recorded, then the earliest first trimester BMI (up to 12 weeks post-LMP) was used.

The Massachusetts General Brigham Institutional Review Board approved all aspects of this study, with a waiver of informed consent as no patient contact was required, the study was considered minimal risk, and consent could not feasibly be obtained. We followed the STROBE reporting guideline for cohort studies.

### Exposure definition

Every birth was linked to a pregnant individual’s SARS-CoV-2 PCR test result at any point in pregnancy, defined as the period between the last menstrual period and the date of birth (within 1 day). The electronic data warehouse (EDW) integrates SARS-CoV-2 PCR results from hospital network laboratories as well as laboratories outside the network if those results are available in the CareEverywhere feature of Epic. Universal screening for SARS-CoV-2 by PCR on admission to Labor and Delivery during the delivery admission was implemented across MGB hospitals from April 2020 through the end of December 2021. During the study period, pregnant individuals with a positive result on a rapid home antigen test were advised to confirm with a hospital-accessible PCR test, per hospital infection control policy; this requirement was relaxed after December 2021. Birthing persons with a positive SARS-CoV-2 PCR test during pregnancy (up to 1 day after delivery) were identified as cases, and birthing persons with a negative SARS-CoV-2 PCR test and no positive SARS-CoV-2 PCR tests were identified as controls. We excluded birthing persons with no SARS-CoV-2 test result at any point during pregnancy.

### Outcome definition

For all included offspring, we extracted ICD-10 billing codes and problem lists. The primary outcome of interest was defined as the presence of at least one diagnosis code associated with obesity, adiposity, or other cardiovascular and metabolic diseases or risk factors within the Healthcare Cost and Utilization Project (HCUP) Clinical Classification Software (CCS) Category 058, “Other nutritional; endocrine; and metabolic disorders.” Codes included in the primary outcome are E66x (Overweight and obesity), E67x (Other hyperalimentation), and E11x (Type 2 Diabetes Mellitus) codes. As in our prior studies (15,16), we also included diagnoses that are not typically assigned in early childhood that are associated with metabolic syndrome in adulthood to create a consistent outcome definition that can be used for long-term follow-up of the cohort in future investigations. A complete list of codes included in the primary outcome is available as Supplementary File 1.

### Offspring anthropometric data

EDW values for offspring weight, length, and BMI were extracted for offspring at 4 time points: birth, 6 months (180 days of life), 12 months (365 days), and 18 months (545 days). At each time point, we selected the closest measurement to the anchor day and within 30 days before and after; if no measurement was recorded within this time frame, no value was reported. Birth measurements were identified as the closest measurement to the date of birth within 7 days. Birthweight-for-gestational age percentiles were calculated using sex-specific growth curves validated for use in preterm and term infants (22). Small for gestational age (SGA) was defined as a birthweight less than the 10^th^ percentile and large for gestational age (LGA) as greater than the 90^th^ percentile. Weight-for-age and length-for-age z-scores at 6-, 12-, and 18-month timepoints were determined using sex-specific 2006 WHO Child Growth Standards (23). For infants born preterm (<37 weeks), the infant’s corrected gestational age (i.e. the child’s age in weeks at the time of measurement minus number of weeks born prior to the estimated date of delivery) was used to calculate the weight-for-age and length-for-age z-score. Z-scores were calculated using the R *peditools* package (v0.3.0.9009). BMI-for-age percentiles populated into medical record flowsheets were extracted directly from the EDW and converted to z-scores. BMI-for-age z-scores were used to classify offspring as at risk for overweight (z ≥1.0 and <2.0) or with overweight/obesity (z ≥2.0). (24).

### Data analysis

We first fit a logistic regression model examining the association between maternal SARS-CoV-2 status during pregnancy and the presence or absence of one or more cardiometabolic outcome diagnostic codes within the first 18 months of life, adjusting for maternal age (years), maternal race and ethnicity, offspring sex, gestational diabetes, and maternal pre-pregnancy BMI, as these factors have been demonstrated to impact offspring growth, development, and cardiometabolic outcomes.(4,25–28) Non-independence of multiple births was addressed by considering observations to be clustered within deliveries; the glm.cluster command in the R *miceadds* package (v3.11-6) was used to generate robust standard errors. We report adjusted estimates of effect and 95% confidence intervals (CI).

To test the hypothesis that SARS-CoV-2 exposure during pregnancy would be associated with differences in fetal and infant growth profiles, we compared differences in anthropometric measures between groups using Welch Two-Sample t-test. These measures included: mean birthweight percentiles, mean weight-for-age z-scores, length-for-age z-scores, and BMI-for-age z-scores between offspring exposed and unexposed to maternal SARS-CoV-2 infection during pregnancy. The proportions of exposed and unexposed offspring categorized as SGA, normal weight, or LGA at birth were compared with Pearson’s Chi-square test, as were proportions of offspring meeting criteria for at risk of overweight or overweight/obesity at 6, 12, and 18 months. All analyses used R 4.0.3 (The R Foundation for Statistical Computing, Vienna, Austria), with statistical significance defined as two-tailed p<0.05.

## Results

### Cohort characteristics

Sociodemographic and clinical information of the cohort is presented in Table 1. The full cohort included 29,510 infants from live births (1,599 SARS-CoV-2-positive cases and 27,911 negative controls; see Supplementary Figure S1 for flow diagram of pandemic cohort derivation). All 29,510 infants had achieved at least 12 months of age at the time of analysis, and there were 20,984 who had achieved at least 18 months of age (1,071 exposed and 19,913 unexposed). Birthing individuals with SARS-CoV-2 were younger (median [IQR]: 31.7 [27.9, 35.2] vs 33.4 [30.4, 36.2] years, p<0.001) and more likely to have public insurance (40% vs 16%, p<0.001). Self-reported race and ethnicity differed between individuals with versus without a SARS-CoV-2 positive test during pregnancy, with a greater proportion of individuals identifying as Hispanic and smaller proportion as White in the SARS-CoV-2 positive group, consistent with previous reports demonstrating disparities in infection risk by race/ethnicity in the greater Boston area and in the US population during the first year of the pandemic.(29,30) The proportion of births occurring preterm did not differ significantly between groups in this cohort (10% in SARS-CoV-2 positive group vs 9% in SARS-CoV-2 negative group). In pregnancies with a positive SARS-CoV-2 test, 54% of infections occurred during the third trimester, 29% in the second trimester, and 17% in the first trimester.

**Table 1.** Sociodemographic and clinical characteristics of individuals with live births who were tested for SARS-CoV-2 during pregnancy (April 1, 2020 – December 31, 2021).

| Characteristic | Maternal SARS-CoV-2 Negative, N = 27,911 | Maternal SARS-CoV-2 Positive, N = 1,599 | p-value |
| --- | --- | --- | --- |
| **Maternal age, years** | 33.4 (30.4, 36.2) | 31.7 (27.9, 35.2) | <0.001 |
| **Maternal race** |  |  | <0.001 |
| White | 19,481 (70%) | 850 (53%) |  |
| Asian | 2,838 (10%) | 76 (4.8%) |  |
| Black or African American | 2,402 (8.6%) | 212 (13%) |  |
| Other | 2,542 (9.1%) | 396 (25%) |  |
| Unknown | 648 (2.3%) | 65 (4.1%) |  |
| **Maternal ethnicity** |  |  | <0.001 |
| Not Hispanic | 23,288 (83%) | 993 (62%) |  |
| Hispanic | 3,879 (14%) | 559 (35%) |  |
| Unknown | 744 (2.7%) | 47 (2.9%) |  |
| **Maternal public insurance** | 4,554 (16%) | 636 (40%) | <0.001 |
| **Delivery hospital type** |  |  | <0.001 |
| Academic medical center | 16,231 (58%) | 1,061 (66%) |  |
| Community hospital | 11,680 (42%) | 538 (34%) |  |
| **Offspring sex** |  |  | 0.8 |
| Female | 13,599 (49%) | 774 (48%) |  |
| Male | 14,310 (51%) | 825 (52%) |  |
| Unknown | 2 | 0 |  |
| **Pre-existing or gestational diabetes** | 3,803 (14%) | 242 (15%) | 0.088 |
| **Maternal pre-pregnancy BMI (kg/m<sup>2</sup>)** | 24.8 (22.1, 29.1) | 26.9 (23.6, 31.3) | <0.001 |
| Unknown | 8,683 | 531 |  |
| **Mode of delivery** |  |  | 0.13 |
| Cesarean section | 9,041 (32%) | 547 (34%) |  |
| Vaginal | 18,870 (68%) | 1,052 (66%) |  |
| **Gestational age (weeks)** | 39.29 (38.29, 40.14) | 39.14 (38.14, 39.89) | <0.001 |
| Unknown | 10 | 3 |  |
| **Preterm birth (<37 weeks)** |  |  | 0.061 |
| Term | 25,501 (91%) | 1,439 (90%) |  |
| Preterm | 2,410 (9%) | 160 (10%) |  |
| **Multiple gestation** | 1,070 (3.8%) | 70 (4.4%) | 0.3 |
| **Trimester of SARS-CoV-2 infection** |  |  |  |
| 1st | N/A | 258 (17%) |  |
| 2nd | N/A | 456 (29%) |  |
| 3rd | N/A | 841 (54%) |  |
| Unknown | N/A | 44 |  |

Continuous data represented as median (IQR) and group differences analyzed by Wilcoxon rank sum test. Categorical data represented as N (%) and group differences analyzed by Pearson's Chi-square test.

### Primary outcome

By 18 months, 72 of 1,071 exposed offspring (6.7%) and 879 of 19,913 unexposed offspring (4.4%) had received a cardiometabolic diagnosis (crude OR 1.47 [95% CI, 1.10-1.94], p=0.007). The individual ICD-10 codes observed in one or more exposed offspring by 18 months are presented in Supplementary Table 1 and most commonly included “Abnormal weight gain” (R63.5), “Overweight” (E66.3) and “BMI pediatric, 85% to less than 95^th^ percentile for age” (Z68.53). In a logistic regression model adjusting for maternal age, race, ethnicity, gestational diabetes, offspring sex, and maternal pre-pregnancy BMI, the adjusted OR of receiving a cardiometabolic diagnosis within 18 months based on pregnancy SARS-CoV-2 exposure status was 1.37 [1.01-1.83]; p=0.04; Figure 1). Secondarily, we examined rates of diagnosis in the larger cohort for whom at least 12 months of follow-up was available and observed a similar pattern. In this group, 77 of 1,599 exposed offspring (4.8%) and 1,010 of 27,911 unexposed offspring (3.6%) had received a cardiometabolic diagnosis (crude OR 1.35 [95% CI, 1.05, 1.70, p=0.014; adjusted OR 1.28 [0.96-1.68; p=0.082]; Supplementary Figure S2).

**Figure 1.**
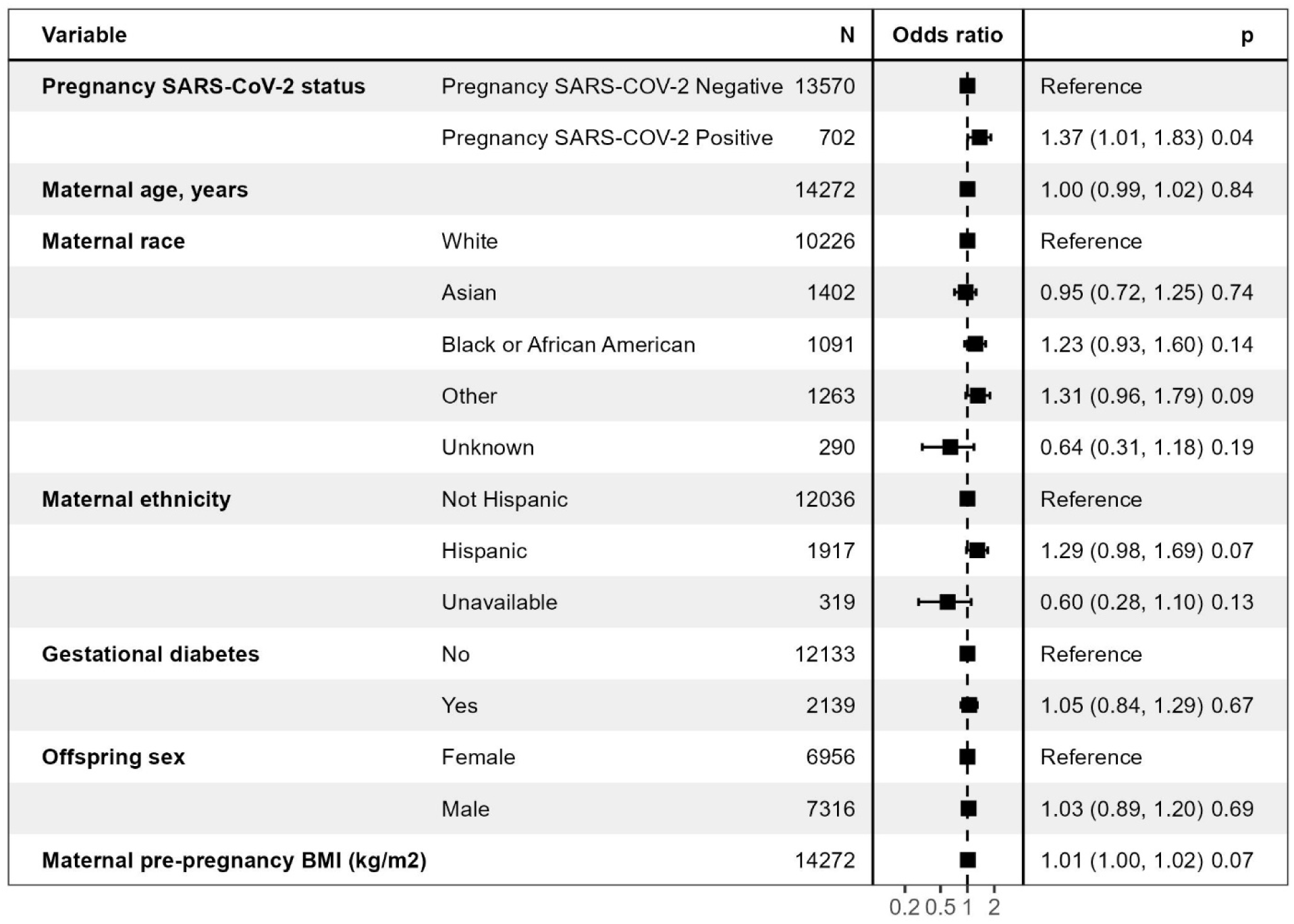
Multiple logistic regression forest plot of cardiometabolic diagnosis in offspring by 18 months of age.

### Offspring weight, length and BMI z-scores at birth and 6, 12, and 18 months

There are conflicting reports linking SARS-CoV-2 to fetal growth restriction (31–33), which in other contexts has been linked to adverse offspring cardiometabolic outcomes (34,35). To examine fetal growth restriction in our cohort, we next compared median gestational age-corrected birthweight percentiles between exposed and unexposed offspring, as well as the proportion of newborns meeting criteria for SGA. Although absolute birthweights were lower in exposed versus unexposed neonates [mean (SD):3252g (568) vs 3303g (562), p<0.001)], there was no difference in gestational age-corrected birthweight percentiles between groups (SARS-CoV-2 positive [mean (SD)]: 49^th^ percentile (26) SARS-CoV-2 negative: 50^th^ percentile (26), nor in rates of SGA (84/1599 (5.3%) vs 1334/27911 (4.8%), p=0.4) between cases and controls (Table 2). The proportion of LGA neonates also did not differ significantly between cases and controls (107/1599 (6.8%) vs 1961/27911 (7.1%), p=0.6).

**Table 2.** Birthweights and gestational age-adjusted birthweight percentiles of exposed and unexposed offspring.

| **Characteristic** | **Pregnancy SARS-CoV-2 Negative, N = 27,911** | **Pregnancy SARS-CoV-2 Positive, N = 1,599** | **p-value** |
| --- | --- | --- | --- |
| **Birthweight (g)** | 3,303 (562) | 3,252 (568) | <0.001 |
| Unknown | 199 | 10 |  |
| **Birthweight category** |  |  | 0.4 |
| Very low birthweight (<1500g) | 250 (0.9%) | 19 (1.2%) |  |
| Low birthweight (<2500g) | 1,678 (6.1%) | 103 (6.5%) |  |
| Normal birthweight | 25,784 (93%) | 1,467 (92%) |  |
| Unknown | 199 | 10 |  |
| **Birthweight percentile for gestational age\*** | 50 (26) | 49 (26) | 0.2 |
| Unknown | 222 | 14 |  |
| **Small for gestational age (SGA)** | 1,334 (4.8%) | 84 (5.3%) | 0.4 |
| Unknown | 222 | 14 |  |
| **Large for gestational age (LGA)** | 1,961 (7.1%) | 107 (6.8%) | 0.6 |
| Unknown | 222 | 14 |  |

Continuous data represented as mean (SD) and differences assessed by Welch's two-sample t-test; categorical as n (%) and differences assessed by Pearson's Chi-squared test. \*Birthweight percentiles using gestational age-specific and sex-specific growth curves validated for use in term and preterm infants (Fenton, 2003). SGA is defined as less than the 10<sup>th</sup> percentile for gestational age. LGA is defined as greater than the 10<sup>th</sup> percentile for gestational age.

To better understand the precise anthropometric changes in offspring that might be associated with observed offspring diagnostic codes, we next examined differences in mean weight, length, and BMI z-scores between exposed and unexposed offspring at 6-month intervals. At birth, mean BMI-for-age z-scores were higher in exposed versus unexposed offspring (p<0.01). At 6 months of age, mean weight-for-age and BMI-for-age z-scores were higher in exposed offspring compared to unexposed controls (Figure 2; Supplementary Table 2). At 6 months of age, the mean difference in BMI-for-age z-score between exposed and unexposed offspring was 0.19 standard deviation units (95% CI: 0.10, 0.29, p<0.001). At 12 and 18 months, weight-for-age and BMI-for-age z-scores were numerically but not statistically significantly higher in exposed offspring versus unexposed controls (Figure 2; Supplementary Table 2).

**Figure 2.**
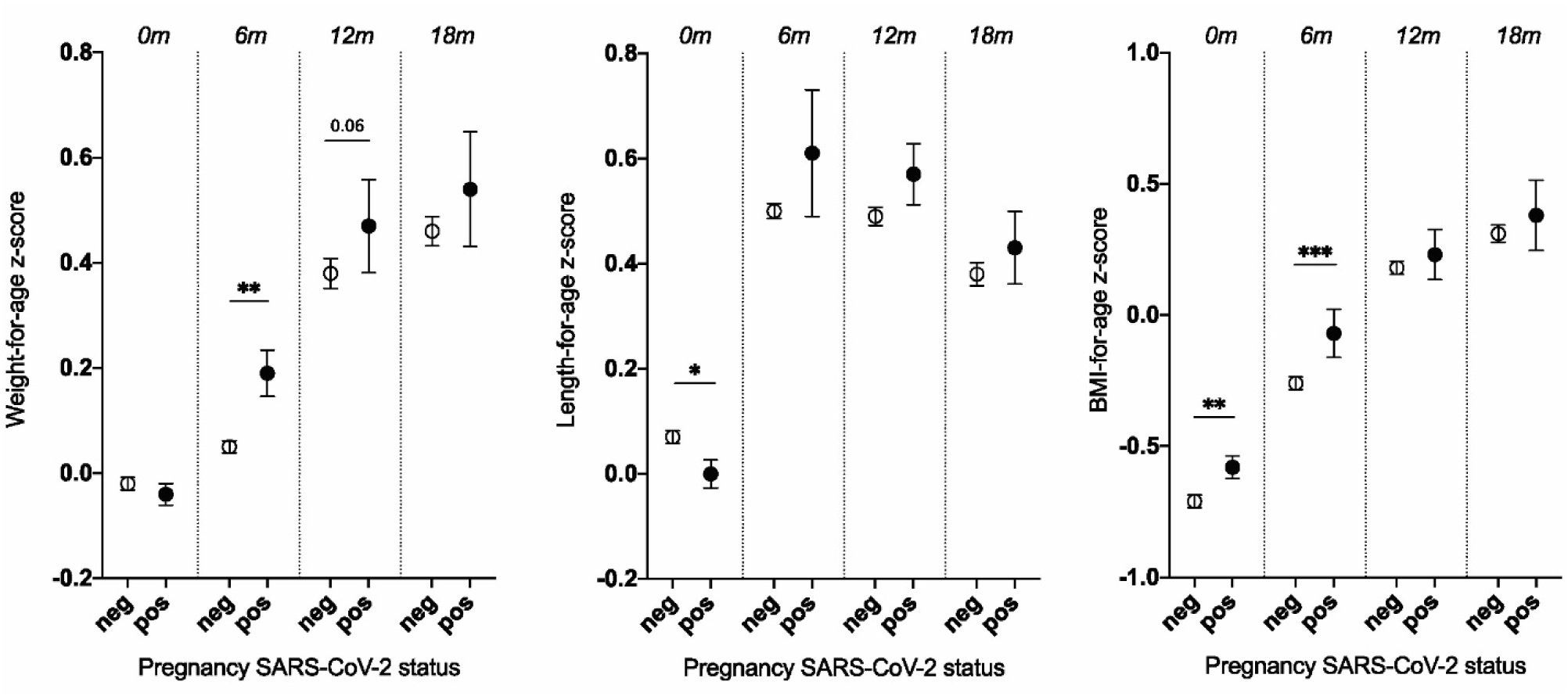
Weight-for-age, length-for-age, and BMI-for-age z-scores in unexposed and exposed offspring. SARS-CoV-2 negative controls are represented by open circles and SARS-CoV-2 positive cases by closed circles at 6-month time intervals. Dot-and-whisker plots depict group mean and SEM. Differences between group means at each time point assessed by Welch’s t-test. *p<0.05, **p<0.01, ***p<0.001. Neg = no SARS-CoV-2 positive test during pregnancy. Pos = SARS-CoV-2 positive test during pregnancy. Z-scores determined sex-specific population norms for children 0-2 years of age (WHO, 2006); corrected gestational age used for offspring born <37 weeks; weight-for-age and length-for age z-score at birth (0m) determined using sex-specific population norms validated for use in term and preterm infants (Fenton 2003).

To assess the clinical relevance of observed differences in mean BMI z-score between groups, we next assessed differences in the proportion of offspring with a BMI z-score of 1.0-1.99 or 2.0 or higher, classifications which have been used to identify children age 0-5 as at risk of overweight, or as having overweight/obesity, respectively (24,36). At 6 months of age, 79 of 480 (16.5%) exposed offspring were classified as at risk of, or meeting criteria for, overweight/obesity, versus 873 of 7161 (12.2%) unexposed offspring (p=0.006). Although similar patterns were noted at 12 and 18 months, differences in the proportion of offspring at risk of, or meeting criteria for, overweight/obesity between groups were not statistically significant at these timepoints. Supplementary Table 3 presents details of BMI z-score categorizations between groups at each time point.

## Discussion

In this EHR-based cohort of 29,510 offspring born during the SARS-CoV-2 pandemic, we found that offspring born to individuals with SARS-CoV-2 infection during pregnancy had significantly greater likelihood of receiving a cardiometabolic diagnosis by the first 18 months of life, with differences emerging within the first year of life. Preceding these diagnoses, we identified significantly higher weight-for-age and BMI-for-age z-scores among exposed versus unexposed offspring at 6 months of age, with a greater proportion of exposed versus unexposed offspring meeting BMI-for-age criteria for overweight or overweight/obesity. BMI-for-age and weight-for-age z-scores were numerically higher at 12 and 18 months, although this finding did not reach statistical significance at those timepoints. Taken together, these results suggest that maternal SARS-CoV-2 exposure during pregnancy is potentially associated with offspring risk for future cardiometabolic disease, observable in early postnatal life.

Maternal SARS-CoV-2 infection during pregnancy has been associated with placental malperfusion (37–39), inflammatory placental signatures (40–42), and fetal growth restriction (40), and thus has the potential for impacting long-term offspring cardiometabolic risk via placental programming (13,34,35). In this large, diverse pandemic-era cohort, we identified that offspring exposed to maternal SARS-CoV-2 infection are at increased risk of receiving a cardiometabolic diagnosis by 18 months, even after adjusting for covariates known to impact offspring cardiometabolic risk, including race/ethnicity, maternal BMI and diabetes. In this context, our findings support the hypothesis that *in utero* exposure to maternal SARS-CoV-2 infection may have an independent effect on offspring cardiometabolic programming. Conversely, overt fetal growth restriction is not likely to play a major role in driving increased cardiometabolic risk in this cohort, as we identified no difference in the birthweight percentiles nor in proportion of SGA infants between exposed and unexposed groups, with only 5.3% of SARS-CoV-2-exposed offspring meeting criteria for SGA. This proportion is consistent with the findings of the COVID-19 Surveillance for Emerging Threats to Mothers and Babies Network (SET-NET) during the first wave of the pandemic, which identified that 5.9% of exposed offspring were SGA (43), and other large cohort studies and meta-analyses identifying no significant risk of SGA with SARS-CoV-2 infection during pregnancy (41,42). Whether the observed higher BMI-for-age at birth in our cohort reflects fetal fat distribution patterns associated with cardiometabolic risk will be important to assess in future prospective cohorts, as newborn body mass composition is not measured during routine clinical care and was thus not able to be assessed in this study.

Differences in the primary outcome between exposed and unexposed offspring were due in part to an increase in diagnostic codes associated with abnormal weight gain and risk for obesity. Consistent with this finding, as well as with our prior work demonstrating significantly greater change in BMI z-scores in the first year of life in exposed offspring (44,45), we observed that both mean postnatal weight-for-age and BMI-for-age z-scores were significantly higher at 6 months of age in exposed offspring. Moreover, the proportion of offspring meeting criteria for being at risk of overweight and as having overweight/obesity was greater in exposed offspring at 6 months of age. The capability of BMI-for-age z-scores in infancy to predict early childhood obesity and adiposity is well-established (43), and strong agreement has been demonstrated between weight-for-length and BMI-for-age values in clinical and research settings (46). Recent work suggests that being classified as overweight or obese by BMI z-score (i.e. greater than or equal to 2) at any time between 6 and 24 months of age is associated with obesity, adiposity, insulin resistance, and metabolic risk in early adolescence (44). The first 6 months of life may be a critical window for intervention to reduce the impact of maternal infections during pregnancy on cardiometabolic programming. Future work should consider the interplay between SARS-CoV-2-associated maternal and placental immune activation during pregnancy and postnatal factors impacting offspring health and development, including breastfeeding, food insecurity, and other social determinants of health.

As a large, retrospective pandemic-era cohort study, this study has several strengths. All live births including those without SARS-CoV-2 exposure occurred during the SARS-CoV-2 pandemic, and the use of contemporaneous controls avoids confounding by unmeasured exposures that might impact cohorts using historical controls. To account for differences in gestational age at birth between groups, we calculated all birthweight percentiles using sex-specific population-level data that have been validated for use in both preterm and term infants (47). Furthermore, gestational age-corrected z-scores were used for assessment of postnatal growth in preterm infants in both groups, reflecting clinical practice patterns and contributing to the external validity of our findings (47).

We also recognize several limitations of this study. Information on other factors that may influence postnatal growth in the first 18 months of life, including maternal breastfeeding status and presence or absence of food insecurity, was not available. Given our findings identifying differences in the primary outcome including codes associated with abnormal weight gain, but not in weight or BMI z-scores at 18 months, it is likely that our study was underpowered to detect small differences in measurements between groups at this time point. Moreover, although effects of viral strain, trimester of exposure, and prior maternal vaccination are of great clinical interest, larger cohorts will be required to achieve sufficient power to examine these effects. Finally, although the Mass General Brigham system represents deliveries occurring at both community and academic hospitals across Eastern Massachusetts, its generalizability to other populations remains to be determined.

In conclusion, these data suggest that SARS-CoV-2 infection during pregnancy is associated with higher offspring weight-for-age and BMI-for-age z-scores at 6 months of age and increased risk of a cardiometabolic diagnosis by 18 months of age, compared to contemporaneous unexposed offspring. The first 6 months of life may therefore represent a critical window for intervention to ameliorate the programming effects of *in utero* SARS-CoV-2 exposure. The intersection of the intrauterine and postnatal environment, including the role of social determinants of health, on offspring cardiometabolic health outcomes will be important to assess in future observational studies.

## Data Availability

All data produced in the present study are available upon reasonable request to the authors.

