## Supplementary for "Offspring cardiometabolic outcomes and postnatal growth trajectories after exposure to maternal SARS-CoV-2 infection"

Supplementary Figures:

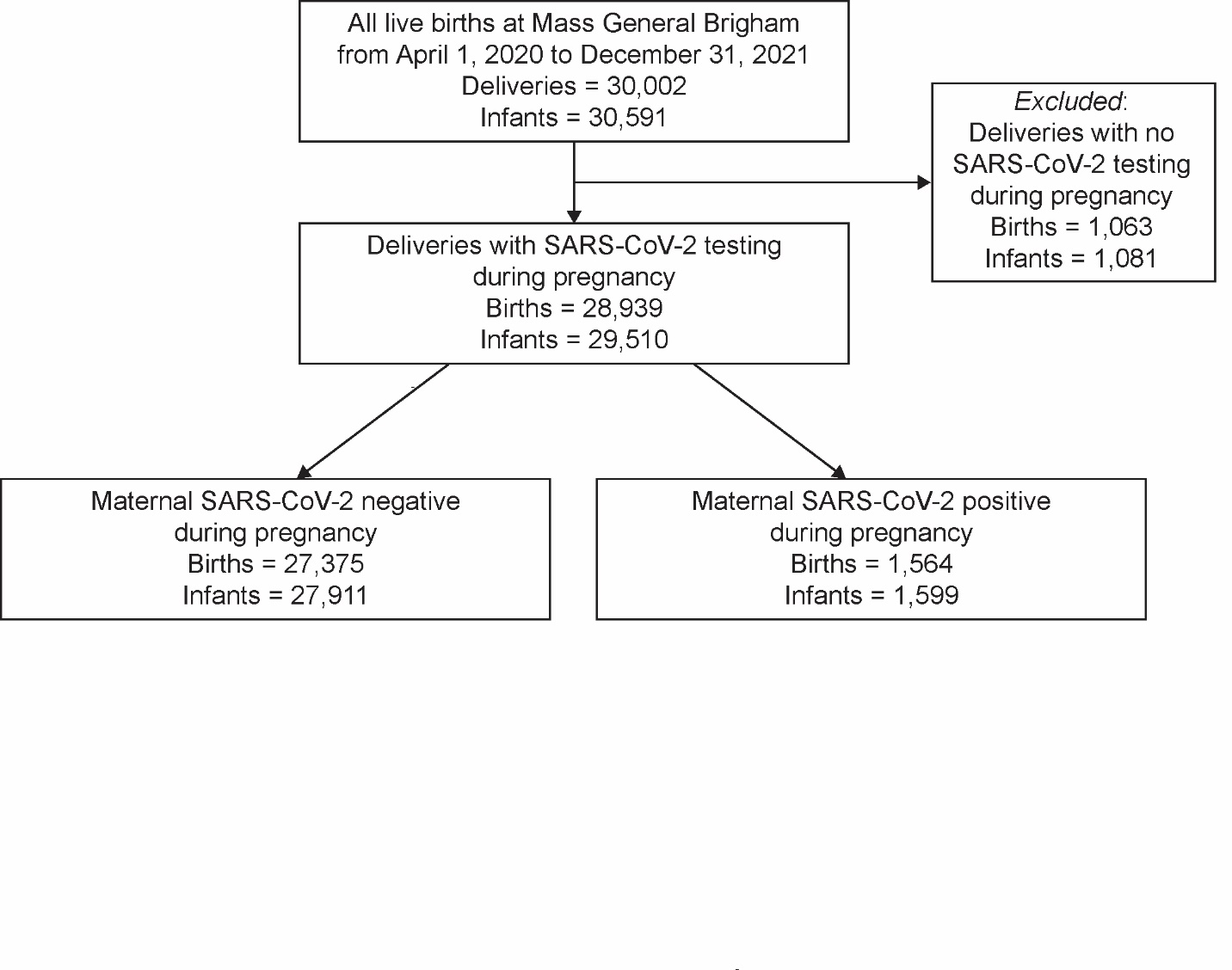

**Supplementary Figure S1**. **Flow diagram of pandemic cohort derivation.**

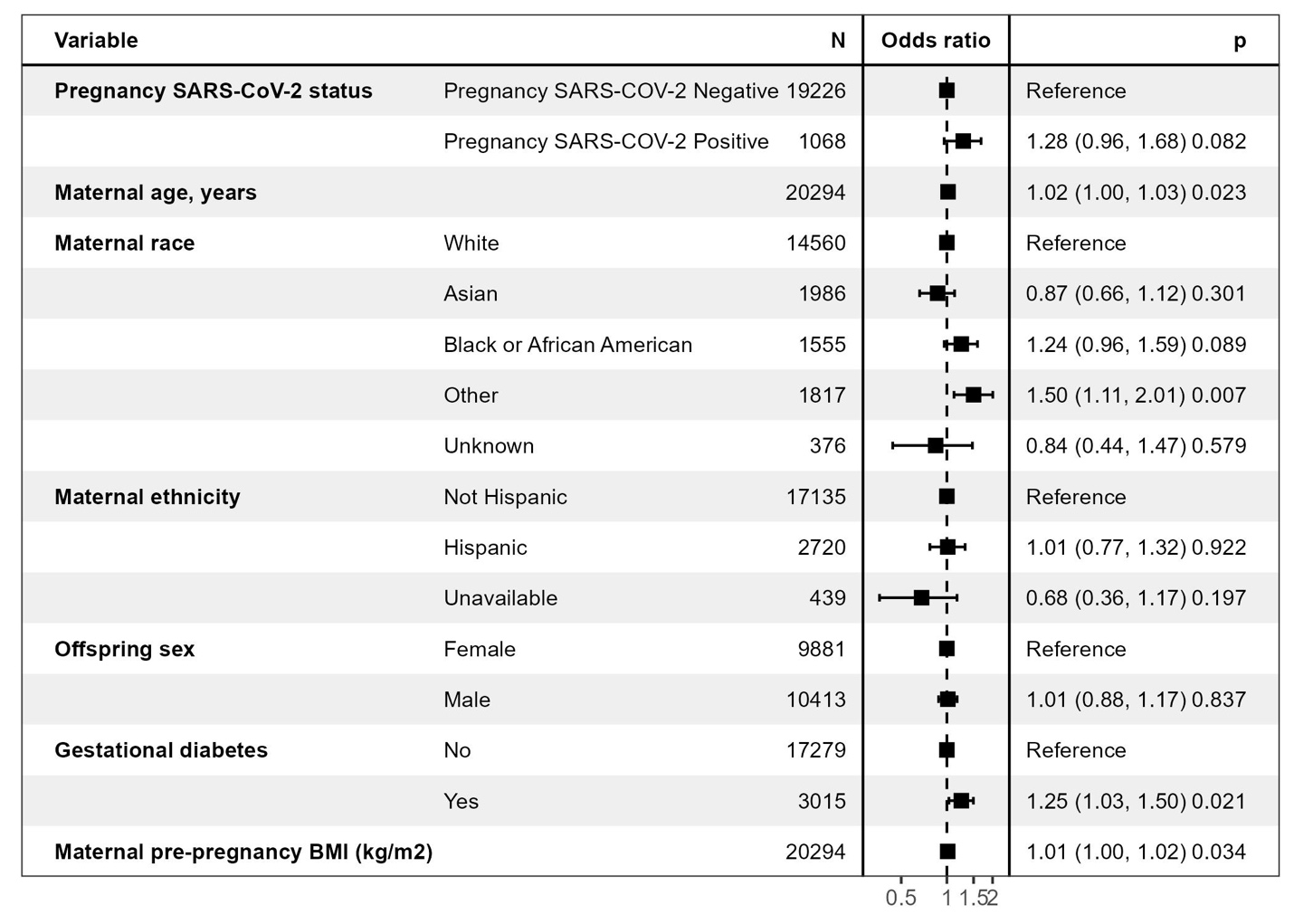

**Supplementary Figure S2.** **Multiple logistic regression forest plot of cardiometabolic diagnosis in offspring by 12 months of age.**

Supplementary Tables:

Supplementary Table 1. Frequency and description of individual cardiometabolic disorder ICD-10-CM codes in offspring cases and controls at 18 months.

| **ICD-10-CM Codes** | **Description** | **Pregnancy SARS-COV-2 Negative (N=879)** | **Pregnancy SARS-COV-2 Positive (N=72)** |
| --- | --- | --- | --- |
| Z71.3 | Dietary counseling and surveillance | 162 | 11 |
| Z68.54 | BMI pediatric, greater than or equal to 95% for age | 37 | <5 |
| Z68.53 | BMI pediatric, 85% to less than 95th percentile for age | 11 | <5 |
| R63.5 | Abnormal weight gain | 236 | 19 |
| R63.2 | Polyphagia | <5 | <5 |
| R62.50 | Unspecified lack of normal physiologic development in childhood | 491 | 44 |
| E66.3 | Overweight | 14 | <5 |

Column N values represent number of offspring with the primary outcome. Column totals exceed number of affected offspring because an individual may have more than 1 diagnosis. Cell counts less than 5 are replaced with <5 to minimize risk of identifiability, per institutional protocol. Table includes diagnosis codes with 1 or greater affected offspring with SARS-CoV-2 pregnancy exposure.

Supplementary Table 2. Offspring weight-for-age, length-for-age, and BMI-for-age z-scores at 6-month time intervals in cases and controls.

| **Characteristic** | **N** | **Pregnancy SARS-CoV-2 Negative***^1^* | **Pregnancy SARS-CoV-2 Positive***^1^* | **Standardized Mean Difference** | | |
| --- | --- | --- | --- | --- | --- | --- |
|  |  |  |  | **Difference** | **95% CI** | **p-value***^2^* |
| Weight-for-age z-score at 0m^3^ | 29,274 | 0.03 (0.84) | 0.00 (0.84) | -0.02 | -0.07, 0.02 | 0.3 |
| Weight-for-age z-score at 6m^4^ | 8,544 | 0.05 (1.03) | 0.19 (1.04) | 0.15 | 0.06, 0.24 | 0.001 |
| Weight-for-age z-score at 12m^4^ | 8,116 | 0.38 (1.26) | 0.47 (1.05) | 0.09 | 0.00, 0.18 | 0.06 |
| Weight-for-age z-score at 18m^4^ | 5,500 | 0.46 (1.03) | 0.54 (1.03) | 0.08 | -0.04, 0.19 | 0.2 |
| Length-for-age z-score at 0m^3^ | 28,522 | 0.03 (1.28) | -0.04 (1.27) | -0.07 | -0.11,0.00 | 0.03 |
| Length-for-age z-score at 6m^4^ | 7,850 | 0.50 (1.22) | 0.61 (2.67) | 0.11 | -0.13, 0.35 | 0.4 |
| Length-for-age z-score at 12m^4^ | 7,111 | 0.49 (1.41) | 0.57 (1.23) | 0.08 | -0.04, 0.19 | 0.2 |
| Length-for-age z-score at 18m^4^ | 4,398 | 0.38 (1.43) | 0.43 (1.11) | 0.05 | -0.09, 0.20 | 0.5 |
| BMI-for-age z-score at 0m^4^ | 6,743 | -0.71 (0.92) | -0.58 (0.90) | 0.13 | 0.04, 0.22 | 0.004 |
| BMI-for-age z-score at 6m^4^ | 7,641 | -0.26 (1.03) | -0.07 (1.02) | 0.19 | 0.10, 0.29 | <0.001 |
| BMI-for-age z-score at 12m^4^ | 6,997 | 0.18 (1.06) | 0.23 (1.03) | 0.06 | -0.04, 0.15 | 0.3 |
| BMI-for-age z-score at 18m^4^ | 4,340 | 0.31 (1.08) | 0.38 (1.08) | 0.07 | -0.07, 0.21 | 0.3 |

Abbreviations: BMI = body mass index. M = months. CI = Confidence Interval.

^1^Mean (SD), ^2^Welch’s t-test. ^3^Z-scores determined using sex-specific population norms validated for use in term and preterm infants (Fenton 2003). ^4^Z-scores determined sex-specific population norms for children 0-2 years of age (WHO, 2006); corrected gestational age used for offspring born <37 weeks.

Supplementary Table 3. Proportion of offspring at risk of overweight and overweight/obesity at 6 – 18 months.

| **Characteristic** | **Pregnancy SARS-CoV-2 Negative** | **Pregnancy SARS-CoV-2 Positive** |
| --- | --- | --- |
| **6 months** | | |
| BMI z-score <1.0,  N (%) | 6288  (87.8) | 401  (83.5) |
| BMI z-score 1.0-1.99,  N (%) | 734  (10.3) | 66  (13.8) |
| BMI z-score ≥2.0,  N (%) | 139  (1.9) | 13  (2.7) |
| **12 months** | | |
| BMI z-score <1.0,  N (%) | 5167  (80.3) | 348  (77.7) |
| BMI z-score 1.0-1.99, N (%) | 1094  (15.6) | 78  (17.4) |
| BMI z-score ≥2.0,  N (%) | 288  (4.1) | 22  (4.9) |
| **18 months** | | |
| BMI z-score <1.0,  N (%) | 3089  (75.5) | 183  (72.9) |
| BMI z-score 1.0-1.99, N (%) | 751  (18.4) | 52  (20.7) |
| BMI z-score ≥2.0,  N (%) | 249  (6.1) | 16  (6.4) |

BMI-for-age z-scores were defined using sex-specific WHO references for age 0-2 years (WHO, 2006). “At risk of overweight” is defined as BMI z-score 1.0-1.99. “Overweight/obesity” is defined as BMI z-score ≥ 2.0 per de Onis, et al.
